# The effectiveness of medical face masks and respirators in reducing SARS-CoV-2 transmission in community settings: a scoping review

**DOI:** 10.1101/2024.10.23.24315907

**Authors:** Constantine I. Vardavas, Valia Marou, Katerina Aslanoglou, Anastasia Manta, Ioanna Lagou, Zinovia Plyta, Jo Leonardi-Bee, Favelle Lamb, Orlando Cenciarelli, Agoritsa Baka

## Abstract

**Background:** During the COVID-19 pandemic, various public health and social measures (PHSM) were implemented with the primary objective of curtailing the transmission of SARS-CoV-2. This review aims to synthesise existing evidence on the effectiveness of medical facemasks and/or respirators (FFP2/KN95/N95) in reducing SARS-CoV-2 transmission/infection in community settings.

**Methods:** A scoping literature review adhering to PRISMA was performed. All relevant study designs within community settings, excluding modelling studies, published between January 2000 and January 2023 and indexed in Medline and Embase were included with no geographical limitation. Studies not specifying facemask/respirator type or not presenting isolated outcomes for specific facemask/respirator types were excluded.

**Results:** Of the 10,185 studies identified, two randomised controlled trials (RCT) and two case-control studies met all inclusion criteria. The largest RCT identified an adjusted prevalence ratio of 0.89 (95%CI: 0.78-1.00) for medical vs. cloth masks. In a smaller RCT, the between-group difference favoured the mask vs no mask group (-0.3 percentage points; 95%CI: -1.2 to 0.4). Within one case-control study, N95/KN95 respirators (aOR 0.17; 95%CI: 0.05-0.64) or medical masks (aOR 0.34; 95%CI: 0.13-0.90) were associated with statistically significant lower adjusted odds of a positive test result compared to no facemask use. A second case-control study associated medical mask use with reduced COVID-19 risk in unadjusted models (OR 0.25; 95% CI: 0.12-0.53) but this effect was not independently associated with infection in multivariable models (aOR 0.61; 95%CI: 0.25-1.49).

**Conclusions:** Limited published evidence exists on the effectiveness of medical facemask use in community settings. Medical masks and respirators (compared to cloth masks) may reduce SARS-CoV-2 transmission, but interpretation requires caution. Mask use in community settings was rarely implemented in isolation to other PHSMs so deciphering whether the effect is solely because of mask-wearing or a combined effect is extremely challenging necessitating additional studies.

## INTRODUCTION

The coronavirus disease 2019 (COVID-19) pandemic, stemming from the emergence of severe acute respiratory syndrome – coronavirus – 2 (SARS-CoV-2), was an unprecedented and far-reaching challenge to public health services, globally. In response to this crisis, various public health and social measures (PHSMs) were implemented to mitigate the transmission of SARS-CoV-2 (1). Among these PHSM, the use of face masks has emerged as a point of interest, given their potential effectiveness in limiting transmission of SARS-CoV-2 in community settings (2). Facemasks differ by type and can include cloth masks (not intended to be used in healthcare settings or by healthcare workers), medical facemasks (a disposable medical device used by healthcare workers to prevent large respiratory droplets and splashes reaching the mouth and nose of the wearer) and respirators (i.e. FFP2, N95, N99 that are designed to protect the wearer from exposure to airborne contaminants) (3, 4). Medical face masks have been advocated as a means to reduce the release of respiratory droplets from asymptomatic, pre-symptomatic, or mildly symptomatic carriers, as well as reducing the inspiration of infectious droplets, thus playing a role in the broader effort to mitigate the transmission of SARS-CoV-2 in community settings (5).

Lessons learned from prior outbreaks and epidemics of respiratory diseases underscore the efficiency of PHSMs in mitigating the transmission of infectious diseases via respiratory droplets and aerosols, such as Middle East Respiratory Syndrome (MERS), SARS and seasonal influenza (6, 7). During the COVID-19 pandemic, public health authorities in general, recommended that persons should wear a face covering and/or a facemask to reduce infection risk in situations where maintaining physical distance from other persons was not feasible.

There is limited evidence in the literature for the effectiveness of face masks outside healthcare settings (2, 8), with previous reviews predominantly focussing on transmission within healthcare settings and not always distinguishing between the type of face mask used (e.g. cloth vs. medical mask vs. respirator). As a result, the effectiveness of medical face or respirators in community settings during the COVID-19 pandemic remains largely unknown.

This scoping review aimed to assess the existing body of evidence regarding the effectiveness of medical face masks and/or respirators (including FFP2, KN95, and N95 respirators), in the context of the public health response to the COVID-19 pandemic within community settings.

## METHODS

This scoping review was conducted adhering to the Preferred Reporting Items for Systematic Reviews and Meta-Analysis (PRISMA) (9) guidelines. The protocol of this review was pre-reviewed by the European Centre for Disease Prevention and Control (ECDC). The protocol was not pre-registered in any database for systematic reviews.

### Inclusion/exclusion criteria and outcomes

Our research question could be summarised in Population (P), Intervention (I), Comparator (C), Outcomes (O), and Setting (S) (PICOS) as follows: (P) people at risk of respiratory virus infection; (I) adhesion to medical face masks and/or respirators use (FFP2, N95, KN95); (C) compared with either no mask-wearing or cloth mask mask-wearing; (O) reduction in the risk of SARS-CoV-2 infection or transmission; and, (S) in community settings (i.e., schools, public transport, religious settings, workplaces-excluding healthcare settings). All relevant study designs within community settings, excluding modelling studies, were included with no geographical limitation imposed, provided they were published in English between January 2000 and January 2023. Studies were excluded if they did not specify the type of face mask or respirator used and did not present isolated outcomes for individual face mask or respirator types.

### Study selection

Peer-reviewed studies were identified through systematic electronic searches using Ovid Medline and Embase. Subject heading terms, free text words relating to the COVID-19 pandemic and the effectiveness of medical face masks and respirators were used to develop a comprehensive list of terms for the search strategy, which is presented in **Supplementary Table 1**. Studies that met the search criteria were evaluated for their relevance. A pilot round of screening was conducted on a random selection of document titles and abstracts between two reviewers. These documents were independently double-screened by the reviewers to empower consistency in screening and identify areas for amendments in the inclusion criteria. A high measure of inter-rater agreement was achieved (percentage agreement > 90%), and hence, the remaining titles were distributed between the two reviewers and screened independently. Any disagreements were thoroughly discussed with a third reviewer. Once all the titles and abstracts had been screened, the full-text documents of the selected studies were retrieved. The retrieved documents were then re-screened based on the information provided in the full-text article. Any disagreements were discussed with the third reviewer. Documents that passed the inclusion criteria based on the full-text screening were included in the review. The manual research and screening of reference lists of review articles were also conducted to include additional relevant studies that were not retrieved through the primary search.

### Data extraction, synthesis and assessment of study quality

The data extracted were related to the study characteristics and metadata (title/ first author’s name/ year of publication), geographical region, setting, and descriptive findings regarding the effectiveness of medical face masks and/or respirators in reducing SARS-CoV-2 transmission in community settings. A data extraction template was created, and data were independently extracted by the two reviewers, with the results discussed in consensus with the third reviewer. We applied a narrative synthesis approach to assess systematically the data and to describe each included study.

The methodological quality of each included study was evaluated independently by two reviewers using the Joanna Briggs Institute (JBI) standardised critical appraisal tool (10) for the appropriate design. Disagreements were resolved with discussion and, when necessary, adjudication by a third reviewer.

## RESULTS

A total of 10,185 studies were identified according to the specified selection criteria from Ovid Medline and Ovid Embase. After removing duplicates, 8,430 studies were screened by title and abstract, out of which 341 studies were assessed for full-text eligibility. Through the assessment of the full texts, 337 studies were excluded due to limited data, ineligible outcome, setting, study design, timeframe, population, or intervention. Consequently, four studies (11-14) were only eligible to be included in this current scoping review, as depicted in the PRISMA flowchart presented in **Figure 1**. Quality appraisal was performed for the studies for which a JBI critical appraisal tool was available (results presented in **Supplementary Table 2**).

**Figure 1.**
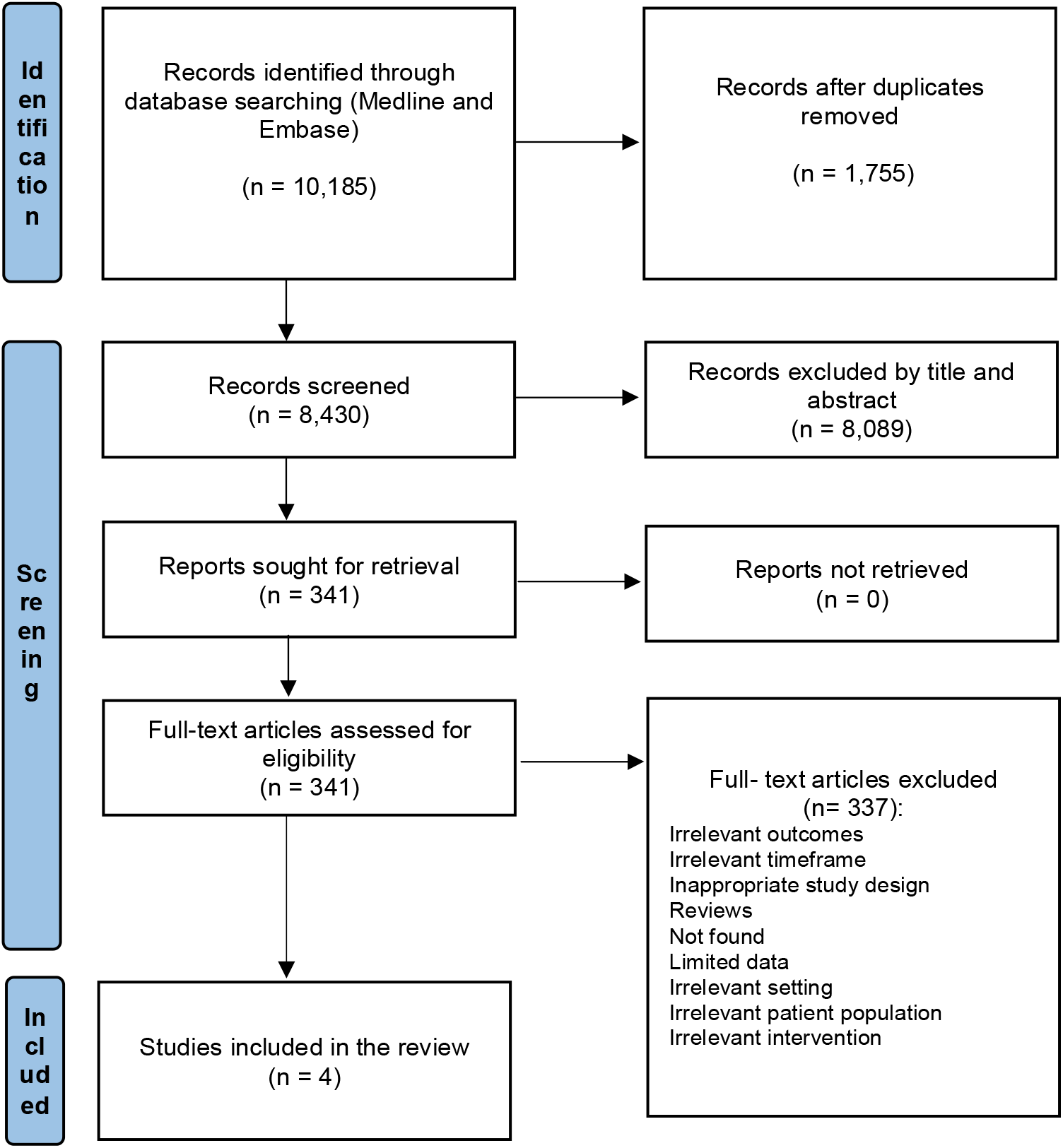
Flowchart of included studies.

### Effectiveness of medical face masks and respirators

**Table 1** presents an overview of the included studies, reporting their setting, study design, participants and the relevant numerical results. Two studies were randomised controlled trials (RCTs) (11, 13) and two were case-control studies (12, 14). In a cluster-randomised trial conducted in rural Bangladesh between November 2020 and April 2021, Abaluck et al. (2021) (11) reported substantial evidence supporting the effectiveness of medical face masks in mitigating symptomatic seroprevalence of SARS-CoV-2. The effects were notably more pronounced and better estimated in communities where medical face masks were distributed compared to those where cloth face masks were provided. Within villages designated to receive medical face masks, they led to a relative reduction in symptomatic seroprevalence of 11.1% [adjusted prevalence ratio = 0.89 (95%CI: 0.78, 1.00); control prevalence = 0.81%; treatment prevalence = 0.72%] compared to cloth face masks. Furthermore, the intervention’s effectiveness in reducing symptomatic seroprevalence was notably higher when medical face masks were used, particularly for the highest-risk demographic segments within the sample (23% reduction for ages 50 to 59, and 35% for ages ≥60). Although cloth masks displayed symptomatic reduction benefits, the impact on symptomatic infections of SARS-CoV-2 is less unequivocal, [adjusted prevalence ratio = 0.94 (95%CI: 0.78, 1.10); control = 0.67%; treatment = 0.61%], contingent on the imputation of missing values for non-consenting adults. It’s worth noting that the number of villages employing cloth face masks (N=100) was half of those utilising medical face masks (N=200), so results should be interpreted with caution. Finally, the intervention led to a reduction in COVID-19-like symptoms under either face mask type (p = 0.000 for medical; p = 0.066 for cloth), but the effect size in medical face mask villages was 30 to 80% larger depending on the specification.

**Table 1.** Overview of included studies and descriptive outcomes regarding the effectiveness of medical facemasks and/or respirators.

| Author et al., | Study design | Countries | Timeframe | Type of facemasks | Setting/Population | Descriptive findings regarding the effectiveness of medical facemasks and/or respirators |
| --- | --- | --- | --- | --- | --- | --- |
| Abaluck et al., 2022 (11) | Cluster randomised controlled trial | Bangladesh | November 2020 - April 2021 | Medical masks vs cloth masks | N =600 villages, N = 342,183 adults | Medical masks lead to a relative reduction in symptomatic seroprevalence of 11.1% (adjusted prevalence ratio = 0.89 [0.78, 1.00]; control prevalence = 0.81%; treatment prevalence = 0.72%) compared to cloth masks.<br>Although cloth masks displayed symptomatic reduction benefits, the impact on symptomatic infections of SARS-CoV-2 is less unequivocal, (adjusted prevalence ratio = 0.94 [0.78, 1.10]; control = 0.67%; treatment = 0.61%)<br>The intervention led to a reduction in COVID-19– like symptoms under either mask type (p = 0.000 for medical; p = 0.066 for cloth), but the effect size in medical mask villages was 30 to 80% larger depending on the specification. |
| Andrejko et al., 2022 (12) | Case control | California, USA | February 18 – December 1, 2021 | N95/KN95 respirators and medical masks | California residents (n=534 with details on type of mask use) | In a sensitivity analysis restricting to participants who did not report known or suspected contact (N = 316), conditional logistic regression models were used to estimate that the unadjusted odds ratios of face mask use by type of face mask with matching strata defined by the week of SARS-CoV-2 testing: 0.13 (95% CI = 0.03–0.61), 0.32 (95% CI = 0.12–0.89), and 0.36 (95% CI = 0.13–1.00) for N95/KN95 respirators, medical masks, or cloth masks, respectively, relative to no face mask or respirator use.<br>Wearing an N95/KN95 respirator (aOR = 0.17; 95% CI = 0.05–0.64) or wearing a medical mask (aOR = 0.34; 95% CI = 0.13–0.90) was associated with lower adjusted odds of a positive test result compared with not wearing a mask (Table 3).<br>Wearing a cloth mask (aOR = 0.44; 95% CI = 0.17–1.17) was associated with lower adjusted odds of a positive test compared with never wearing a face covering but was not statistically significant. |
| Bundgaard et al., 2020 (13) | Randomised controlled trial (DANMASK-19) | Denmark | 3 April to 2 June 2020 | Medical masks | Adults spending more than 3 hours per day outside the home who are not recommended to wearing face masks at work (n=6024) | In an intention-to-treat analysis, the between group difference was - 0.3 percentage point (CI, - 1.2 to 0.4 percentage point; P= 0.38) (odds ratio [OR], 0.82 [CI, 0.54 to 1.23]; P= 0.33) in favor of the mask group<br>When this analysis was repeated with multiple imputation for missing data due to loss to follow-up, it yielded similar results (OR, 0.81 [CI, 0.53 to 1.23]; P = 0.32).<br>Excluding participants in the mask group who reported nonadherence (7%), SARS-CoV-2 infection occurred in 40 participants (1.8%) in the mask group and 53 (2.1%) in the control group (between-group difference, - 0.4 percentage point [CI, - 1.2 to 0.5 percentage point]; P= 0.40) (OR, 0.84 [CI, 0.55 to 1.26]; P= 0.40). |
| Doung-Ngern et al., 2020 (14) | Case control | Thailand | March - May, 2020 | Medical masks | COVID-19 cases and noninfected controls from contact tracing records of the central SRRT team | The consistent use of masks throughout the exposure period to individuals infected with SARS-CoV-2 exhibited a distinct correlation with a lower infection risk.<br>In unadjusted models medical masks were associated with a reduced risk of COVID-19 with a crude OR of 0.25 (95% C.I: 0.12–0.53) compared to no mask use however, because of collinearity with mask-wearing compliance, it was not included in the final model |
|  |  |  |  |  | at the Department of Disease Control (DDC), MoPH, Thailand. | Mask type in a separate multivariable model was not independently associated with infection ($p = 0.54$), with an aOR of 0.61 (95%CI: 0.25–1.49) for medical mask vs no mask use.<br>The study identified no substantial evidence of effect modification between mask type and the degree of compliance with mask-wearing |

Bundgaard et al. (2021) (13) conducted an RCT (named DANMASK-19) to assess the effectiveness of incorporating a face mask recommendation as an additional preventive measure against SARS-CoV-2 infection in a community setting, where use of a face mask was infrequent. Participants in the intervention group were provided with medical face masks and eligibility required a minimum of three hours of exposure to individuals outside their homes. In an intention-to-treat analysis, the between-group difference was – 0.3 percentage points (95%CI: -1.2 to 0.4 percentage point; p= 0.38) (OR 0.82; 95%CI: 0.54 – 1.23; p=0.33) in favour of the medical face mask group. When this analysis was repeated with multiple imputations for missing data due to loss to follow-up, it yielded similar results (OR 0.81; 95%CI: 0.53 – 1.23; p=0.32). Excluding participants in the medical face mask group who reported nonadherence (7%), SARS-CoV-2 infection occurred in 1.8% in the medical face mask group and 2.1% in the control group (between-group difference, - 0.4 percentage point, 95%CI: - 1.2 to 0.5 percentage point; p= 0.40) (OR 0.84; 95%CI: 0.55 – 1.26; p=0.40). Despite the non-statistically significant findings, the study provides insights into the anticipated level of protection that medical face mask wearers might expect in a community setting where face mask usage is limited and other PHSM, including physical distancing, are in effect.

Two case-control studies were also identified. Andrejko et al. (2022) (12) performed a test-negative design case-control study which identified that among individuals surveyed, those who reported wearing regularly N95/KN95 respirators exhibited the lowest adjusted odds of infection, followed closely by those wearing medical face masks. Among 534 participants who specified the type of face covering they typically used, wearing N95/KN95 respirators (aOR 0.17; 95%CI: 0.05–0.64) or medical face masks (aOR 0.34; 95%CI: 0.13– 0.90) was associated with statistically significant lower adjusted odds of a positive test result compared with not wearing any face mask/respirator. Wearing a cloth facemask (aOR 0.44; 95%CI: 0.17–1.17) was associated with lower adjusted odds of a positive test compared with never wearing a face covering but was not statistically significant.

The second case-control study conducted by Doung-ngern et al. (2020) (14) examined the effectiveness of personal protective measures in mitigating SARS-CoV-2 infection. In unadjusted models, medical face masks were associated with a reduced risk of SARS-CoV-2 infection with a crude OR of 0.25 (95% CI: 0.12–0.53) compared to no face mask use. However, because of collinearity with medical face mask-wearing adherence, mask type was not included in the final model. In a separate multivariable model, face mask type was not independently associated with infection (aOR 0.61; 95%CI: 0.25–1.49, p=0.54) for medical face mask vs. no face mask use. Furthermore, the study identified no substantial evidence of effect modification between face mask type and the degree of compliance with face mask-wearing. However, within this study we cannot exclude the possibility of recall bias where participants may have misclassified respirators as medical masks.

## DISCUSSION

Only limited evidence was identified on the effectiveness of medical face masks and respirators within community settings. Among the limited number of studies identified, the largest RCT indicated a protective effect of medical vs. cloth face mask use (11), while the DANMASK-19 study indicated a protective, albeit not statistically significant role of medical face mask use in reducing SARS-CoV-2 transmission in community settings (13). The two case-control studies both suggested a protective effect of respirators and medical face masks compared to no face mask use, however in multivariable models mask use indicated collinearity with face mask warning compliance, while no evidence of effect modification by face mask type was noted (12, 14).

Our findings support the existing epidemiological data from both observational studies and RCTs that have assessed the effectiveness of medical face masks and/or respirators in reducing transmission within hospital settings (15) for other respiratory viruses (16). In a meta-analysis by Chu et al., medical face mask use was in general associated with a larger reduction in risk of infection from SARS-CoV-2, SARS-CoV, and MERS-CoV while the authors also noted that respirators may be associated with more protection from viral infection than medical face masks or cloth face masks (6). Similarly, Kim et al. (7) also noted that N95 respirators were the most effective (OR 0.30; 95% CI, 0.20–0.44 ; GRADE, low) in providing protection against coronavirus infections (SARS, MERS, SARS-CoV-2) consistently across subgroup analyses of causative viruses and clinical settings (7), while medical masks also indicated a potentially protective effect (OR, 0.72; 95% CI, 0.51–1.01). However, it is important to note that while SARS-CoV-2, SARS-CoV, and MERS-CoV are all coronaviruses, the characteristics of the diseases, including their reproduction number, caused by these viruses are substantially different (17). Moreover, it is important to note that there is substantial experimental evidence of the comparative effectiveness of different face mask types, however these have been implemented in lab settings or are based on modelling studies (18-27), not within community settings.

Only a small number of studies have looked at the possible effects of the implementation of other PHSMs implemented in-parallel to face mask use including physical distancing, stay-home orders, business closures, and mass gathering restrictions, all of which impact SARS-CoV-2 transmission and add further complexity to the evaluations. Furthermore, when these parameters are documented, they result in mostly ecological associations, with limited or no information available on how effectively people adhere to these PHSMs. Given these limitations, it is challenging to separate the comparative effectiveness of PHSMs and draw firm conclusions. Moreover, interventions around proper facemask use may also have impacted the results in the two RCTs identified, as participants also received information on proper mask use, which may have impacted the results. Results from the largest RCT we identified suggest that combining face mask provision, role-modelling, and the promotion of facemask use, rather than mask distribution and role-modelling alone, was instrumental to achieving the full effect (11).

Our scoping review’s strength lies in its systematic examination of the literature and comprehensive evaluation of evidence regarding the effectiveness of medical face masks and respirators in community settings, while its focus on SARS-CoV-2 and not on previous respiratory virus outbreaks (MERS-CoV, SARS-CoV, Influenza) adds to the study’s novelty. Nevertheless, it is essential to acknowledge some limitations. Firstly, the specificity of our research question, focusing solely on medical face masks and respirators in community settings, restricted substantially the number of eligible studies for inclusion. This resulted in a small pool of data for our analysis, which prevented us from conducting a meaningful meta-analysis. Hence, we opted for a qualitative synthesis approach to descriptively present the selected studies’ findings. Another limitation is the potential for publication bias. As studies with positive or significant results tend to be published more frequently, this may possibly lead to an overestimation of medical face mask and respirator effectiveness in community settings. Furthermore, it is impossible to rule out the possibility of recall or response bias of respondents within the original studies that have been included in our review. The presence of small strata further hindered the differentiation between types of face masks or variations in face mask use across different settings and therefore, wider confidence intervals and non-significance were observed for certain estimates that nonetheless suggested potential protective effects. Moreover, we cannot exclude the possibility of bias in the original studies (recall bias, social desirability bias, misclassification bias etc). Finally, the studies did not consider other behaviours and population factors that could have influenced infection risk and mask use including PHSM (28, 29, 30) and conflicting messaging that was noted during the pandemic, which may have prevented the public from adhering to the recommended PHSM(s). Addressing potential factors that reduce compliance and community engagement activities and training on the appropriate use of medical face masks in a community setting are recommended within the context of respiratory pandemics (31, 32).

Despite these limitations, our scoping review provides valuable insights into the effectiveness of medical face masks and respirators in community settings and indicates the necessity for the provision of additional observational and experimental studies, including the implementation of RCTs, as studies have indicated the superiority of respirators and medical face masks vs. cloth masks within experimental lab settings (27). With regards to the types of studies needed to elucidate these research questions from a community perspective, longitudinal studies with individual-level data could be an appropriate study design as are individual and community cluster RCTs, as also case-control studies that could provide important information about risk estimates at an individual level, while controlling for the other relevant risk factors at both regional- and individual levels, providing information on compliance, type of face mask used and other in parallel PHSMs that may contribute to the variability currently identified.

## CONCLUSIONS

There is limited published evidence on the effectiveness of medical face mask and respirator use in community settings, which, to the extent identified, provide supportive evidence that medical face masks and respirators (compared to cloth face masks) may reduce SARS-CoV-2 transmission. However, due to a significant number of confounding factors and limitations in study design, including collinearity with adherence to appropriate face mask use recommendations, proper face mask use and other PHSMs implemented in parallel limit the generalisability of the findings. The findings from this scoping review add to the evidence base on the effectiveness of face mask use and identify key methodological aspects that should be considered in study development, which ultimately could be used to inform public health strategies, shape policy decisions, and guide future research aimed at optimising face mask usage as a PHSM during potential future respiratory pandemics.

## Data Availability

Data sharing is not applicable to this article as no new data were created or analysed in this study.

## Acknowledgements

We would like to thank the archivists, Katerina Papathanasaki and Stella Vogiatzidaki for their assistance in data archiving and full text retrieval.

## Ethics Statement

Not applicable.

## REFERENCES

1. Lai S, Ruktanonchai NW, Zhou L, Prosper O, Luo W, Floyd JR, et al. Effect of non-pharmaceutical interventions to contain COVID-19 in China. Nature. 2020;585(7825):410–3.

2. Coclite D, Napoletano A, Gianola S, del Monaco A, D’Angelo D, Fauci A, et al. Face Mask Use in the Community for Reducing the Spread of COVID-19: A Systematic Review. Frontiers in Medicine. 2021;7.

3. European Committee for Standardization. Respiratory protective devices Brussels, Belgium [Available from: https://standards.cen.eu/dyn/www/f?p=204:110:0::::FSP_PROJECT,FSP_ORG_ID:32928,6062&cs=1FC98AD34A5EE26A0CB5A6155ED4D6E5.

4. ECDC. Considerations for the use of face masks in the community in the context of the SARS-CoV-2 Omicron variant of concern. Stockholm; 2022.

5. ECDC. Using face masks in the community: first update. Stockholm: European Centre for Disease Prevention and Control; 2021.

6. Chu DK, Akl EA, Duda S, Solo K, Yaacoub S, Schünemann HJ. Physical distancing, face masks, and eye protection to prevent person-to-person transmission of SARS-CoV-2 and COVID-19: a systematic review and meta-analysis. Lancet. 2020;395(10242):1973–87.

7. Kim MS, Seong D, Li H, Chung SK, Park Y, Lee M, et al. Comparative effectiveness of N95, surgical or medical, and non-medical facemasks in protection against respiratory virus infection: A systematic review and network meta-analysis. Rev Med Virol. 2022;32(5):e2336.

8. Leech G, Rogers-Smith C, Monrad JT, Sandbrink JB, Snodin B, Zinkov R, et al. Mask wearing in community settings reduces SARS-CoV-2 transmission. Proc Natl Acad Sci U S A. 2022;119(23):e2119266119.

9. Page MJ, McKenzie JE, Bossuyt PM, Boutron I, Hoffmann TC, Mulrow CD, et al. The PRISMA 2020 statement: an updated guideline for reporting systematic reviews. BMJ. 2021;372:n71.

10. Moola S, Munn Z, Tufanaru C, Aromataris E, Sears K, Sfetc R, et al. Chapter 7: Systematic Reviews of Etiology and Risk. 2020.

11. Abaluck J, Kwong LH, Styczynski A, Haque A, Kabir MA, Bates-Jefferys E, et al. Impact of community masking on COVID-19: A cluster-randomized trial in Bangladesh. Science. 2022;375(6577):eabi9069.

12. Andrejko KL, Pry JM, Myers JF, Fukui N, DeGuzman JL, Openshaw J, et al. Effectiveness of Face Mask or Respirator Use in Indoor Public Settings for Prevention of SARS-CoV-2 Infection - California, February-December 2021. MMWR Morb Mortal Wkly Rep. 2022;71(6):212–6.

13. Bundgaard H, Bundgaard JS, Raaschou-Pedersen DET, von Buchwald C, Todsen T, Norsk JB, et al. Effectiveness of Adding a Mask Recommendation to Other Public Health Measures to Prevent SARS-CoV-2 Infection in Danish Mask Wearers. Annals of Internal Medicine. 2020;174(3):335–43.

14. Doung-Ngern P, Suphanchaimat R, Panjangampatthana A, Janekrongtham C, Ruampoom D, Daochaeng N, et al. Case-Control Study of Use of Personal Protective Measures and Risk for SARS-CoV 2 Infection, Thailand. Emerg Infect Dis. 2020;26(11):2607–16.

15. Bartoszko JJ, Farooqi MAM, Alhazzani W, Loeb M. Medical masks vs N95 respirators for preventing COVID-19 in healthcare workers: A systematic review and meta-analysis of randomized trials. Influenza and Other Respiratory Viruses. 2020;14(4):365–73.

16. Abboah-Offei M, Salifu Y, Adewale B, Bayuo J, Ofosu-Poku R, Opare-Lokko EBA. A rapid review of the use of face mask in preventing the spread of COVID-19. International Journal of Nursing Studies Advances. 2021;3:100013.

17. Jüni P, da Costa BR, Bobos P, Bodmer NS, McGeer A. Revisiting the evidence for physical distancing, face masks, and eye protection. Lancet. 2021;398(10301):663.

18. Arav Y, Klausner Z, Fattal E. Theoretical investigation of pre-symptomatic SARS-CoV-2 person-to-person transmission in households. Sci Rep. 2021;11(1):14488.

19. Asadi S, Cappa CD, Barreda S, Wexler AS, Bouvier NM, Ristenpart WD. Efficacy of masks and face coverings in controlling outward aerosol particle emission from expiratory activities. Sci Rep. 2020;10(1):15665.

20. Bennett JS, Mahmoud S, Dietrich W, Jones B, Hosni M. Evaluating vacant middle seats and masks as Coronavirus exposure reduction strategies in aircraft cabins using particle tracer experiments and computational fluid dynamics simulations. Eng Rep. 2022:e12582.

21. Catching A, Capponi S, Yeh MT, Bianco S, Andino R. Examining the interplay between face mask usage, asymptomatic transmission, and social distancing on the spread of COVID-19. Sci Rep. 2021;11(1):15998.

22. Gurbaxani BM, Hill AN, Paul P, Prasad PV, Slayton RB. Evaluation of different types of face masks to limit the spread of SARS-CoV-2: a modeling study. Scientific Reports. 2022;12(1):8630.

23. Henriques A, Mounet N, Aleixo L, Elson P, Devine J, Azzopardi G, et al. Modelling airborne transmission of SARS-CoV-2 using CARA: risk assessment for enclosed spaces. Interface Focus. 2022;12(2):20210076.

24. Koh XQ, Sng A, Chee JY, Sadovoy A, Luo P, Daniel D. Outward and inward protection efficiencies of different mask designs for different respiratory activities. Journal of Aerosol Science. 2022;160:105905.

25. Lasser J, Sorger J, Richter L, Thurner S, Schmid D, Klimek P. Assessing the impact of SARS-CoV-2 prevention measures in Austrian schools using agent-based simulations and cluster tracing data. Nat Commun. 2022;13(1):554.

26. Mello VM, Eller CM, Salvio AL, Nascimento FF, Figueiredo CM, Silva E, et al. Effectiveness of face masks in blocking the transmission of SARS-CoV-2: A preliminary evaluation of masks used by SARS-CoV-2-infected individuals. PLoS One. 2022;17(2):e0264389.

27. Robinson JF, Rios de Anda I, Moore FJ, Gregson FKA, Reid JP, Husain L, et al. How effective are face coverings in reducing transmission of COVID-19? Aerosol Science and Technology. 2022;56(6):473–87.

28. Agaku I, Nkosi L, Agaku QD, Gwar J, Tsafa T. Factors related to adherence to public COVID-19 prevention behaviors, United States, April–July 2021. Population Medicine. 2022;4(September):1–13.

29. Rajan S, McKee M, Hernández-Quevedo C, Karanikolos M, Richardson E, Webb E, et al. What have European countries done to prevent the spread of COVID-19? Lessons from the COVID-19 Health system response monitor. Health Policy. 2022;126(5):355–61.

30. Seale H, Dyer CEF, Abdi I, Rahman KM, Sun Y, Qureshi MO, et al. Improving the impact of non-pharmaceutical interventions during COVID-19: examining the factors that influence engagement and the impact on individuals. BMC Infect Dis. 2020;20(1):607.

31. Brainard J, Jones NR, Lake IR, Hooper L, Hunter PR. Community use of face masks and similar barriers to prevent respiratory illness such as COVID-19: a rapid scoping review. Euro Surveill. 2020;25(49).

32. Afolabi AA, Ilesanmi OS. Community engagement for COVID-19 prevention and control: A systematic review. Public Health Toxicol. 2022;2(2):1–17.

