## Supplementary Tables 2,3 for "The effectiveness of medical face masks and respirators in reducing SARS-CoV-2 transmission in community settings: a scoping review"

**Supplementary Table 2. Quality Appraisal of Case-control studies (n=2)**

| Questions | Andrejko et al., 2022 | Doung-Ngern et al., 2020 |
| --- | --- | --- |
| 1. Were the groups comparable other than the presence of disease in cases or the absence of disease in controls? | YES | YES |
| 2. Were cases and controls matched appropriately? | YES | YES |
| 3. Were the same criteria used for identification of cases and controls? | YES | YES |
| 4. Was exposure measured in a standard, valid and reliable way? | YES | YES |
| 5. Was exposure measured in the same way for cases and controls? | YES | YES |
| 6. Were confounding factors identified? | YES | N/A |
| 7. Were strategies to deal with confounding factors stated? | YES | N/A |
| 8. Were outcomes assessed in a standard, valid and reliable way for cases and controls? | YES | YES |
| 9. Was the exposure period of interest long enough to be meaningful? | YES | YES |
| 10. Was appropriate statistical analysis used? | YES | YES |
| Rating | 10/10 | 10/10 |

**Supplementary Table 3. Quality Appraisal of Randomized Control Trials (RCTs) (n=2)**

| Questions | Abaluck, et al., 2021 | Bundgaard et al., 2021 |
| --- | --- | --- |
| 1. Was true randomization used for assignment of participants to treatment groups? | YES | YES |
| 2. Was allocation to treatment groups concealed? | YES | YES |
| 3. Were treatment groups similar at the baseline? | YES | YES |
| 4. Were participants blind to treatment assignment? | YES | YES |
| 5. Were those delivering the treatment blind to treatment assignment? | YES | YES |
| 6. Were treatment groups treated identically other than the intervention of interest? | YES | YES |
| 7. Were outcome assessors blind to treatment assignment? | YES | YES |
| 8. Were outcomes measured in the same way for treatment groups? | YES | YES |
| 9. Were outcomes measured in a reliable way? | YES | YES |
| 10. Was follow up complete and if not, were differences between groups in terms of their follow up adequately described and analysed? | YES | YES |
| 11. Were participants analysed in the groups to which they were randomized? | YES | YES |
| 12. Was appropriate statistical analysis used? | YES | YES |
| 13. Was the trial design appropriate and any deviations from the standard RCT design (individual randomization, parallel groups) accounted for in the conduct and analysis of the trial? | YES | YES |
| Rating | 13/13 | 13/13 |
