## Supplementary Table 1 for "The effectiveness of medical face masks and respirators in reducing SARS-CoV-2 transmission in community settings: a scoping review"

**Supplementary Table 1. Search strategy**

| **Database: Ovid MEDLINE(R) ALL <1946 to April 11, 2023>** |  |
| --- | --- |
| **KEY TERMS** | **HITS** |

| 1 | *coronavirus/ or *Coronavirus disease 2019/ or *coronavirus infections/ | 215811 |
| --- | --- | --- |
| 2 | *SARS-CoV-2/ or *COVID-19/ | 176469 |
| 3 | (COVID19 or COVID-19 or SARS-CoV-2).ti,ab. | 325358 |
| 4 | (("2019" adj (novel or new) adj corona*) or ("2019" adj (CoV or nCoV)) or (coronavirus adj (disease adj "2019")) or ((Novel or New) adj Corona*) or (SARS adj2 (coronaviridae or coronavirus)) or ((sars or Coronavirus) adj "2") or 2019ncov).ti,ab. | 86707 |
| 5 | or/1-4 | 350485 |
| 6 | (N95 or FFP2 or P2 or "Korea 1st class" or "filtering mask*" or "filtering respirator*").ti,ab,kf. | 39520 |
| 7 | 6 and | 51445 |
| 8 | limit 7 to yr="2020 -Current" | 1420 |

| **Database: Embase ALL <1946 to 2023 April 11>** |  |
| --- | --- |
| **KEY TERMS** | **HITS** |

| 1 | Coronavirinae/ or coronavirus disease 2019/ or Coronaviridae Infections/ or Coronaviridae/ | 349490 |
| --- | --- | --- |
| 2 | *SARS-CoV-2/ or *COVID-19/ or Severe acute respiratory syndrome coronavirus 2/ | 221087 |
| 3 | (COVID19 or COVID-19 or SARS-CoV-2).ti,ab. | 395852 |
| 4 | (("2019" adj (novel or new) adj corona*) or ("2019" adj (CoV or nCoV)) or (coronavirus adj (disease adj "2019")) or ((Novel or New) adj Corona*) or (SARS adj2 (coronaviridae or coronavirus)) or ((sars or Coronavirus) adj "2") or 2019ncov).ti,ab. | 97998 |
| 5 | or/1-4 | 447750 |
| 6 | (N95 or FFP2 or P2 or "Korea 1st class" or "filtering mask*" or "filtering respirator*").ti,ab,kf. | 48223 |
| 7 | 6 and | 51782 |
| 8 | limit 7 to yr="2020 -Current" | 1766 |

| **Database: Ovid MEDLINE(R) ALL <1946 to May 15, 2023>** |  |
| --- | --- |
| **KEY TERMS** | **HITS** |

| 1 | *coronavirus/ or *Coronavirus disease 2019/ or *coronavirus infections/ | 220505 |
| --- | --- | --- |
| 2 | *SARS-CoV-2/ or *COVID-19/ | 181147 |
| 3 | (COVID19 or COVID-19 or SARS-CoV-2).ti,ab. | 331979 |
| 4 | (("2019" adj (novel or new) adj corona*) or ("2019" adj (CoV or nCoV)) or (coronavirus adj (disease adj "2019")) or ((Novel or New) adj Corona*) or (SARS adj2 (coronaviridae or coronavirus)) or ((sars or Coronavirus) adj "2") or 2019ncov).ti,ab. | 88089 |
| 5 | or/1-4 | 357388 |
| 6 | Masks/ | 7258 |
| 7 | (Mask? or facemask? or face-mask? or N95 or FFP2 or P2 or "Korea 1st class" or "filtering mask*" or "filtering respirator*").ti,ab,kf. | 90439 |
| 8 | 6 or 7 | 91975 |
| 9 | 5 and 8 | 9090 |
| 10 | limit 9 to yr="2020 -Current" | 9039 |

| **Database: Embase ALL <1946 to 2023 May 15>** |  |
| --- | --- |
| **KEY TERMS** | **HITS** |

| 1 | Coronavirinae/ or coronavirus disease 2019/ or Coronaviridae Infections/ or Coronaviridae/ | 358305 |
| --- | --- | --- |
| 2 | *SARS-CoV-2/ or *COVID-19/ or Severe acute respiratory syndrome coronavirus 2/ | 230894 |
| 3 | (COVID19 or COVID-19 or SARS-CoV-2).ti,ab. | 404764 |
| 4 | (("2019" adj (novel or new) adj corona*) or ("2019" adj (CoV or nCoV)) or (coronavirus adj (disease adj "2019")) or ((Novel or New) adj Corona*) or (SARS adj2 (coronaviridae or coronavirus)) or ((sars or Coronavirus) adj "2") or 2019ncov).ti,ab. | 99746 |
| 5 | or/1-4 | 458044 |
| 6 | Masks/ | 10492 |
| 7 | (Mask? or facemask? or face-mask? or N95 or FFP2 or P2 or "Korea 1st class" or "filtering mask*" or "filtering respirator*").ti,ab,kf. | 114184 |
| 8 | 6 or 7 | 116080 |
| 9 | 5 and 8 | 10838 |
| 10 | limit 9 to yr="2020 -Current" | 10798 |
